# Socioeconomic position, socioenvironmental conditions, and subjective cognitive decline among midlife and older LGBTQIA+ adults in California

**DOI:** 10.64898/2026.11.11.26367750

**Authors:** Nguyen K. Tran, Ethan Cicero, Nicole Rosendale, Gowri Sunder, Carol Dawson-Rose, Annesa Flentje

## Abstract

**Objectives:** To examine which measure of socioeconomic position and socioenvironmental conditions has the strongest relationship with subjective cognitive decline (SCD) among midlife and older lesbian, gay, bisexual, transgender, queer, intersex, asexual, and other sexual and gender minority (LGBTQIA+) adults.

**Methods:** We conducted a cross-sectional analysis of 3869 LGBTQIA+ adults aged 50 years or older participating in a community-engaged survey in California. Using logistic regression, we examined overall associations between each indicator of socioeconomic position and socioenvironmental conditions and SCD. We used a dominance analysis to assess the relative importance of each indicator based on the proportion of variance in SCD it explained.

**Results:** Overall, 14.7% of the sample reported SCD. Midlife and older LGBTQIA+ adults in a lower socioeconomic position (*i.e.,* lower education, unemployment, lower household income, fewer assets, financial insecurity, food insecurity, and unstable housing) had higher odds of SCD than those in a higher position, with significant odds ratios (OR) ranging from 1.28 to 2.55. In contrast, perceiving one’s community as safe for LGBTQIA+ people was associated with lower odds of SCD (OR 0.47; 95% CI 0.38, 0.58). Dominance analysis indicated that household assets was the most important factor, explaining 4.4% of the variance in SCD (95% CI 3.5%, 5.4%), followed by household income (2.9%; 95% CI 2.2%, 3.8%) and perceived community safety (2.2%; 95% CI 1.4%, 3.1%).

**Discussion:** Supporting the cognitive health of LGBTQIA+ people may require attention to both financial security across the life course and the broader social environments.

## INTRODUCTION

Subjective cognitive decline (SCD) is characterized by self-reported experiences of worsening or frequent confusion or memory loss (Jessen et al., 2014). While not all individuals who report SCD will progress on to develop cognitive decline, it remains an important early indicator associated with an elevated risk of mild cognitive impairment and Alzheimer’s disease and related dementias (ADRD) (Jessen et al., 2014; Rabin et al., 2017; Rostamzadeh et al., 2022). SCD can also interfere with individuals’ capacity to perform activities of daily living, thereby compromising their quality of life (Hill et al., 2017). Because interventions to delay or prevent ADRD may be most effective at the earliest stages of disease progression, there is scientific and public health interest in identifying factors that contribute to SCD (Jessen et al., 2020; Wong et al., 2025).

Approximately 11% of U.S. adults aged 45 years or older report SCD (Taylor et al., 2018). This burden has been shown to be disproportionately pronounced among lesbian, gay, bisexual, queer, intersex, asexual, and other sexual and gender minority (LGBTQIA+) populations (Flatt et al., 2021; Fredriksen-Goldsen et al., 2022; Huo et al., 2024). According to the minority stress model, higher rates of physical health problems experienced by LGBTQIA+ people are a result of distinct and additive stressors beyond those experienced by the general population, such as experiences with rejection, expectation of stigma, and internalized negativity (Flentje et al., 2020; Hendricks & Testa, 2012; Meyer, 2003). Among LGBTQIA+ adults aged 45 years or older, 15.7% reported SCD, a prevalence 50% higher than their non-LGBTQIA+ counterparts (Flatt et al., 2021). Further compounded by the intersection of gender and race, midlife and older transgender adults, particularly those from minoritized ethnoracial communities, reported having 1.7 to 2.5 times greater odds of SCD compared to their White cisgender counterparts (Cicero et al., 2023).

A substantial body of research has identified social and economic factors as upstream determinants of cognitive aging and ADRD risk in the general population (Glymour & Manly, 2008; Lövdén et al., 2020; Luo et al., 2025; Rosso et al., 2016; Singh-Manoux et al., 2005; Adkins-Jackson et al., 2023), yet their role within LGBTQIA+ populations remains poorly understood. Existing studies among LGBTQIA+ populations have primarily focused on social isolation and depression as key contributors to SCD (Flatt et al., 2018; Fredriksen-Goldsen et al., 2018; Hsieh et al., 2021; Kim et al., 2025). However, LGBTQIA+ adults also face elevated prevalence of poverty and economic insecurity and are more likely to experience employment loss, food insecurity, and inadequate financial resources to support daily living compared with non-LGBTQIA+ people (Fredriksen-Goldsen et al., 2013; Nguyen et al., 2024). Occupation-based socioeconomic position, such as precarious employment and loss of employment, has been associated with greater depressive symptoms in LGBTQIA+ adults (Kinitz, Tran, Shahidi, et al., 2026; Kinitz, Tran, Soltani, et al., 2026), a proximal factor for SCD (Correro II & Nielson, 2020; Flatt et al., 2018; Hsieh et al., 2021). These conditions may become increasingly consequential for LGBTQIA+ people as they age and underscore the need to better address the social and economic needs that support healthy cognitive aging in this population.

In this study, we aimed to characterize the prevalence of SCD and its relationship to respondents’ socioeconomic position and socioenvironmental conditions in a community-based sample of midlife and older LGBTQIA+ adults in California. We focused on California given its rapidly growing population of both older adults and LGBTQIA+ people (Johnson, 2024; Johnson et al., 2025), and because addressing economic insecurity is a key priority among these groups (University of California, San Francisco, 2024). We then aim to quantify the relative contribution of each social and economic factor that shaped SCD among midlife and older LGBTQIA+ adults to understand which factors may be among the most important in relation to SCD.

## METHODS

### Study Design and Participants

This study is a secondary analysis of data collected through an online cross-sectional survey conducted from January to March 2024 to assess the health, well-being, and service needs of midlife and older LGBTQIA+ adults living in California (University of California, San Francisco, 2024). Participants were recruited using convenience sampling through a variety of outreach strategies led by Openhouse, a nonprofit organization that supports older LGBTQIA+ people. These strategies included the creation of a statewide coalition of LGBTQIA+ service providers, distribution of flyers in community spaces, social media posts, newsletters, a dedicated website hosted by the California Department of Aging, and word-of-mouth among community members, senior service organizations, and events. Additional details regarding community outreach are described elsewhere (University of California, San Francisco, 2024).

The eligibility criteria for the study were self-identifying as LGBTQIA+, being 50 years of age or older, and residing in California including community-dwelling older adults and those in senior living facilities. The survey was administered online through Qualtrics (Qualtrics LLC, Provo, UT) and was available in Chinese, English, Spanish, and Tagalog, the most widely spoken languages in California. Eligible participants were asked to provide informed consent for themselves, and those who completed the survey could choose to enter a drawing to win a $25 gift card. The study was approved by the [redacted] Institutional Review Board.

Several steps were taken to ensure the quality of survey responses during data collection. These included fraud and bot detection tools embedded in Qualtrics and manual reviews of potentially invalid responses. Those identified as likely fraudulent or bot-generated (n = 406) or flagged as invalid during manual review (n = 2) were excluded from the dataset. In addition, we excluded 168 participants with missing outcome data for the current analysis.

### Measures

#### Subjective Cognitive Decline

Subjective cognitive decline was assessed with a single self-reported question based on the Behavioral Risk Factor Surveillance System Cognitive Decline module, “During the past 12 months, have you experienced confusion or memory loss that is happening more often or is getting worse?” Response options included “yes”, “no”, and “I don’t know/I’m not sure”. Those who responded affirmatively were considered to have experienced SCD.

#### Socioeconomic and Socioenvironmental Conditions

We included nine indicators that captured participants’ socioeconomic position and socioenvironmental condition. 1) Education level was categorized into four groups: high school diploma/GED or less, some college, 4-year college degree, and graduate degree. 2) Unemployment was dichotomized as a binary measure. Participants were classified as unemployed if they did not report any form of employment (employed, working 1-39 hours per week; employed, working 40 or more hours per week; self-employed; or temporarily employed) and were not retired. All others were considered unemployed. 3) Household income was categorized into five groups ranging from $30,000 or less to more than $120,000. 4) Household assets were assessed by asking participants to estimate the total value of all assets owned by their household. Responses were grouped into four categories from less than $10,000 to more than $500,000. 5) Financial insecurity was assessed by asking whether participants had struggled to pay bills due to insufficient income in the past 12 months (yes or no). 6) Food insecurity was measured by asking how often participants worried about having enough money for nutritious meals in the past 12 months (yes or no). 7) Housing stability was assessed based on participants’ current housing situation and categorized as stable if they reported having a steady place to live and unstable if they reported being worried about losing housing in the future or not having a steady place to live (*e.g.*, in a shelter, living outside on the street). 8) Transportation was assessed by asking participants to select the methods they typically use to get where they need to go. Participants who selected public transportation were categorized as relying on it; all others were categorized as not relying on public transportation. 9) LGBTQIA+ community safety was assessed through two questions asking how safe participants perceived their current community to be for 1) transgender, genderqueer, and non-binary individuals (gender minority safety) and 2) lesbian, gay, asexual, bisexual, and queer individuals (sexual minority safety), using a 5-point scale (1=extremely safe to 5=extremely unsafe). We then combined these responses to create a composite measure with four categories: safe for both gender and sexual minority people, safe for gender minority people only, safe for sexual minority people only, and unsafe for both groups.

#### Participant Characteristics

Participants self-reported their ethnoracial identity, current gender identity, current sexual orientation, intersex status, place of birth, prior military service, and health insurance. Participants could select multiple options for their ethnoracial identity, gender, and sexual orientation. Sexual orientations were combined to create non-mutually exclusive groups: asexual, bisexual, gay/lesbian, queer, straight, and another sexual orientation. Gender identities were cross stratified with sex assigned at birth to create mutually exclusive groups: cisgender men, cisgender women, gender diverse individuals assigned female at birth, gender diverse individuals assigned male at birth, transgender men, and transgender women. Urban residence (yes or no) was based on self-reported ZIP code. Age groups (50-59, 60-69, ≥70 years) was based on self-reported birthdate and categorized prior to data release to support de-identification. HIV status was dichotomized as living with HIV for those reported “positive” and not living with HIV if otherwise.

#### Mental Health and Substance Use

Mental health measures included loneliness assessed using the UCLA 3-item Loneliness Scale (UCLA-3, range: 3-12) (Hughes et al., 2004), depressive symptoms in the past two weeks assessed using the Patient Health Questionnaire-2 (PHQ-2, range: 0-6) (Kroenke et al., 2003), and PTSD symptoms in the past month assessed using the 6-item PTSD Checklist (PCL-6, range: 6-30) (Lang & Stein, 2005). For all three scales, higher scores indicate greater symptom severity. Substance use included current cigarette smoking and alcohol consumption, with the latter assessed using the Alcohol Use Disorders Identification Test-Consumption (AUDIT-C, range: 0-12), where higher scores indicate greater alcohol consumption and potential hazardous drinking (Bush et al., 1998).

### Statistical Analysis

#### Descriptive analysis

We summarized the socioeconomic and socioenvironmental conditions, sociodemographic characteristics, health conditions, and substance use for the overall sample and by SCD. Differences were assessed using t-test for continuous variables and chi-square or Fisher’s Exact test for categorical variables. Correlations were calculated between each socioeconomic and socioenvironmental indicator.

#### Regression model specification

To evaluate associations between socioeconomic and socioenvironmental conditions and SCD, we fitted separate logistic regression models to estimate the unadjusted and adjusted odds ratios (OR) and associated 95% confidence intervals (CI). We used a directed acyclic graph (**Supplemental Figure 1**) to identify potential confounders and mediators based on the published literature of correlates of cognitive decline and ADRD risk in both the general population and LGBTQIA+ people (Adkins-Jackson et al., 2023; Alzheimer’s Association, 2025; Correro II & Nielson, 2020; Flatt et al., 2018; Fredriksen-Goldsen et al., 2013, 2018; Hsieh et al., 2021; Kim et al., 2025; Livingston et al., 2020; Lövdén et al., 2020; Luo et al., 2025). For our main analysis, we considered the hypothetical timing of covariates in relation individuals’ socioeconomic and socioenvironmental conditions from a life course perspective in order to identify a minimally sufficient set of confounders for adjustment. Adjusted models accounted for age groups, gender groups, ethnoracial groups, US birth, urban residence, prior military service, and living with HIV. While mental health symptoms and substance use may be important confounders, in the context of our study, we have conceptualized them as potential mediators based on prior literature and our hypothesized framework and therefore did not adjust for them in the main analysis (Zwieten et al., 2022). As a sensitivity analysis, we assessed the robustness of the estimated associations in the main analysis to additional adjustment for mental health symptoms (loneliness, depressive symptoms, PTSD symptoms) and substance use (current smoker and alcohol use).

#### Handling of missing data

Approximately 18% of the study sample had any missing data, with non-response rates ranging from 0.1% to 8.7% across all variables. Thus, we addressed missingness using multiple imputation by chained equations, using the *mice* R package to generate 20 imputed datasets (van Buuren & Groothuis-Oudshoorn, 2011). Analyses for our main models were conducted on each imputed dataset, and the results were combined using Rubin’s rules to obtain overall estimates and their standard errors (Rubin, 1978).

#### Dominance analysis

To calculate the relative importance of each socioeconomic and socioenvironmental condition on SCD in LGBTQIA+ midlife and older adults, we used an extension of the dominance analysis procedure that allows for the use of logistic regression (Azen & Traxel, 2009). This approach assesses the change in variance for all possible subset models, allowing assessment of the relative importance of each predictor both alone and in the presence of all other combinations of predictors — that is, how much variance in the observed outcome can be attributed to one factor versus another. However, unlike linear models that use the traditional *R^2^*, we used *pseudo-R^2^*measures, specifically McFadden’s *R^2^*, to determine relative importance. We opted for McFadden’s *R^2^* due to its desirable statistical properties (Azen & Traxel, 2009), including boundness (values vary between 0 to 1), linear invariance (remains unchanged to linear transformation), monotonicity (value does not decrease when variables are added to the model), and intuitive interpretability.

We first fit a multivariable logistic model that included all socioeconomic and socioenvironmental factors while controlling for potential confounders as outlined in our main analysis, representing the total variance explained by our model. Because the dominance analysis software does not support multiply imputed data, we used complete cases for this analysis. We also refitted the same model with imputed data to assess the robustness of the results to the assumed missing data mechanism. We then used the *dominanceanalysis* R package to decompose the total variance and evaluate the relative importance of each socioeconomic and environmental factors while keeping potential confounders constant across all models (Navarrete & Soares, 2025). A bootstrap procedure with 1000 resamples was used to obtain general dominance values (i.e., proportion of variance explained for a given variable averaged across all model sizes) and the corresponding 95% percentile CI. We report general dominance for our analysis to identify which socioeconomic and environmental factors that, on average, contribute most to explaining the variance in SCD among LGBTIQIA+ midlife and older adults. We also reran the dominance analysis to determine whether the relative importance of each socioeconomic and socioenvironmental condition would change after accounting for mental health symptoms and substance use. All analysis were conducted in R version 4.5.1.

## RESULTS

### Descriptive Analysis

The final sample included 3,869 midlife and older LGBTQIA+ respondents (**Table 1**), of whom 34.0% (n=1317) were aged ≥70 years, 11.2% (n=424) were transgender and gender diverse, 21.8% (n=844) reported at least 1 minoritized ethnoracial identity, 86.3% (n=3340) identified as gay or lesbian, and 1.0% (n=40) were intersex. Educational levels were high in the study sample; 73% (n=3813) held at least a 4-year degree, including 42% (n=1624) with graduate degrees **(Table 2**). However, unemployment was at 56.6% (n=2189). Income distribution varied widely, with 32.2% (n=1246) reporting household incomes of >$120,000, while 16.9% (n=655) earned ≤$30,000. Similarly, 41.3% (n=1597) reported household assets of >$500,000, contrasting with 13% (n=502) having <$10,000 in assets. Respondents reported various forms of insecurity: 25.9% (n=1001) experienced financial insecurity, 34.9% (n=1352) faced food insecurity, and 12.9% (n=500) dealt with housing instability. Additionally, 34.0% (n=1314) relied on public transit for their transportation needs. Half (51.3%, n=1986) of respondents felt their community was safe for both sexual and gender minority people, while 20.8% (n=803) perceived it as unsafe.

**Table 1.**
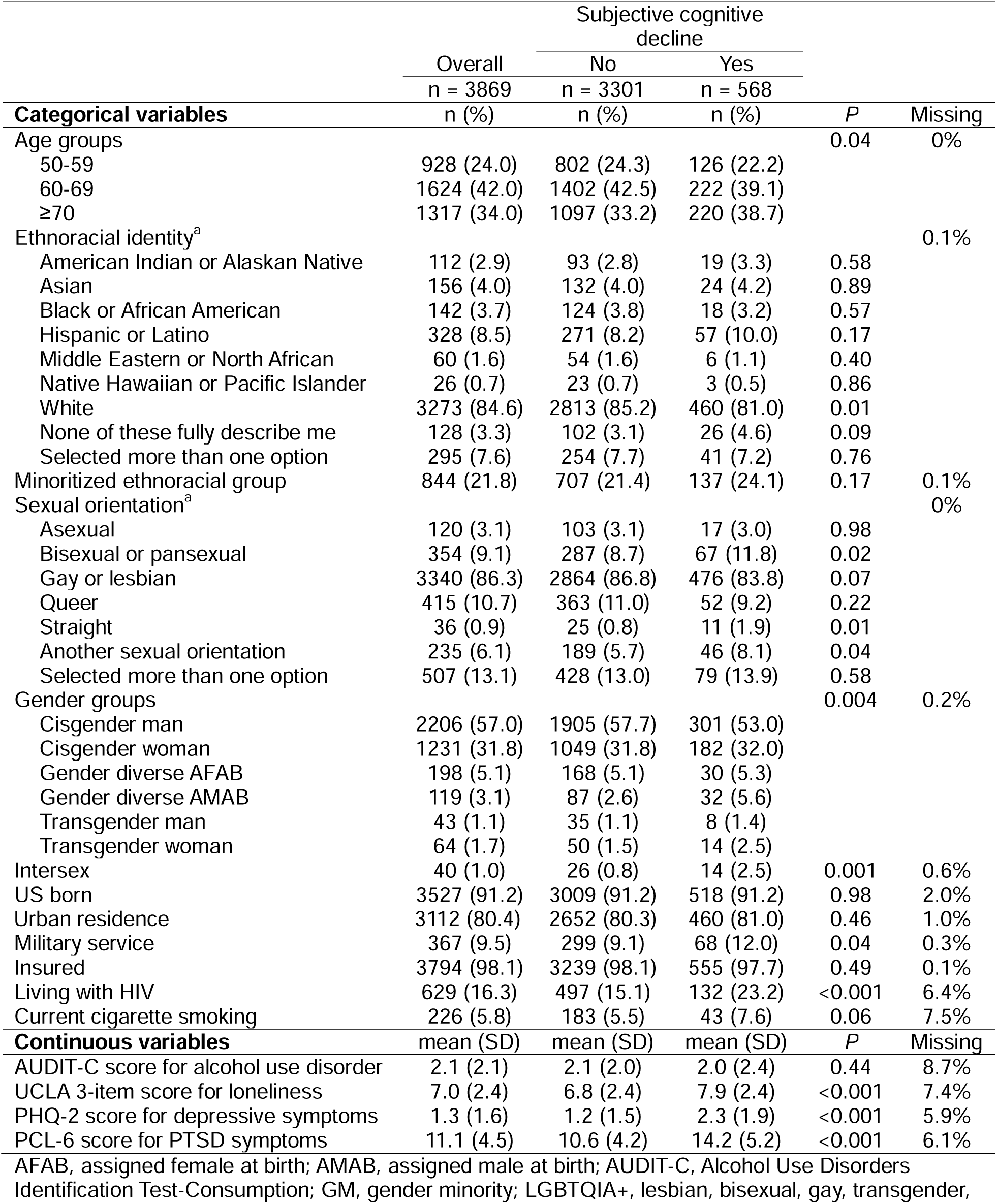
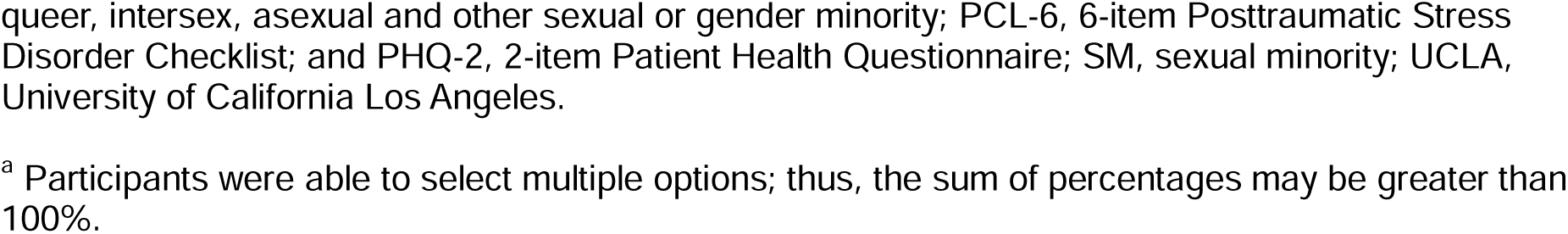
Participant characteristics by subjective cognitive decline among LGBTQIA+ adults aged 50 years or older in California.

|  | Subjective cognitive decline |  |  |  |  |
|---|---|---|---|---|---|
|  | Overall | No | Yes |  |  |
|  | n = 3869 | n = 3301 | n = 568 |  |  |
| **Categorical variables** | n (%) | n (%) | n (%) | *P* | Missing |
| Age groups |  |  |  | 0.04 | 0% |
| 50-59 | 928 (24.0) | 802 (24.3) | 126 (22.2) |  |  |
| 60-69 | 1624 (42.0) | 1402 (42.5) | 222 (39.1) |  |  |
| ≥70 | 1317 (34.0) | 1097 (33.2) | 220 (38.7) |  |  |
| Ethnoracial identity<sup>a</sup> |  |  |  |  | 0.1% |
| American Indian or Alaskan Native | 112 (2.9) | 93 (2.8) | 19 (3.3) | 0.58 |  |
| Asian | 156 (4.0) | 132 (4.0) | 24 (4.2) | 0.89 |  |
| Black or African American | 142 (3.7) | 124 (3.8) | 18 (3.2) | 0.57 |  |
| Hispanic or Latino | 328 (8.5) | 271 (8.2) | 57 (10.0) | 0.17 |  |
| Middle Eastern or North African | 60 (1.6) | 54 (1.6) | 6 (1.1) | 0.40 |  |
| Native Hawaiian or Pacific Islander | 26 (0.7) | 23 (0.7) | 3 (0.5) | 0.86 |  |
| White | 3273 (84.6) | 2813 (85.2) | 460 (81.0) | 0.01 |  |
| None of these fully describe me | 128 (3.3) | 102 (3.1) | 26 (4.6) | 0.09 |  |
| Selected more than one option | 295 (7.6) | 254 (7.7) | 41 (7.2) | 0.76 |  |
| Minoritized ethnoracial group | 844 (21.8) | 707 (21.4) | 137 (24.1) | 0.17 | 0.1% |
| Sexual orientation<sup>a</sup> |  |  |  |  | 0% |
| Asexual | 120 (3.1) | 103 (3.1) | 17 (3.0) | 0.98 |  |
| Bisexual or pansexual | 354 (9.1) | 287 (8.7) | 67 (11.8) | 0.02 |  |
| Gay or lesbian | 3340 (86.3) | 2864 (86.8) | 476 (83.8) | 0.07 |  |
| Queer | 415 (10.7) | 363 (11.0) | 52 (9.2) | 0.22 |  |
| Straight | 36 (0.9) | 25 (0.8) | 11 (1.9) | 0.01 |  |
| Another sexual orientation | 235 (6.1) | 189 (5.7) | 46 (8.1) | 0.04 |  |
| Selected more than one option | 507 (13.1) | 428 (13.0) | 79 (13.9) | 0.58 |  |
| Gender groups |  |  |  | 0.004 | 0.2% |
| Cisgender man | 2206 (57.0) | 1905 (57.7) | 301 (53.0) |  |  |
| Cisgender woman | 1231 (31.8) | 1049 (31.8) | 182 (32.0) |  |  |
| Gender diverse AFAB | 198 (5.1) | 168 (5.1) | 30 (5.3) |  |  |
| Gender diverse AMAB | 119 (3.1) | 87 (2.6) | 32 (5.6) |  |  |
| Transgender man | 43 (1.1) | 35 (1.1) | 8 (1.4) |  |  |
| Transgender woman | 64 (1.7) | 50 (1.5) | 14 (2.5) |  |  |
| Intersex | 40 (1.0) | 26 (0.8) | 14 (2.5) | 0.001 | 0.6% |
| US born | 3527 (91.2) | 3009 (91.2) | 518 (91.2) | 0.98 | 2.0% |
| Urban residence | 3112 (80.4) | 2652 (80.3) | 460 (81.0) | 0.46 | 1.0% |
| Military service | 367 (9.5) | 299 (9.1) | 68 (12.0) | 0.04 | 0.3% |
| Insured | 3794 (98.1) | 3239 (98.1) | 555 (97.7) | 0.49 | 0.1% |
| Living with HIV | 629 (16.3) | 497 (15.1) | 132 (23.2) | <0.001 | 6.4% |
| Current cigarette smoking | 226 (5.8) | 183 (5.5) | 43 (7.6) | 0.06 | 7.5% |
| **Continuous variables** | mean (SD) | mean (SD) | mean (SD) | *P* | Missing |
| AUDIT-C score for alcohol use disorder | 2.1 (2.1) | 2.1 (2.0) | 2.0 (2.4) | 0.44 | 8.7% |
| UCLA 3-item score for loneliness | 7.0 (2.4) | 6.8 (2.4) | 7.9 (2.4) | <0.001 | 7.4% |
| PHQ-2 score for depressive symptoms | 1.3 (1.6) | 1.2 (1.5) | 2.3 (1.9) | <0.001 | 5.9% |
| PCL-6 score for PTSD symptoms | 11.1 (4.5) | 10.6 (4.2) | 14.2 (5.2) | <0.001 | 6.1% |

AFAB, assigned female at birth; AMAB, assigned male at birth; AUDIT-C, Alcohol Use Disorders Identification Test-Consumption; GM, gender minority; LGBTQIA+, lesbian, bisexual, gay, transgender, queer, intersex, asexual and other sexual or gender minority; PCL-6, 6-item Posttraumatic Stress Disorder Checklist; and PHQ-2, 2-item Patient Health Questionnaire; SM, sexual minority; UCLA, University of California Los Angeles.
<sup>a</sup> Participants were able to select multiple options; thus, the sum of percentages may be greater than 100%.

**Table 2.** Socioeconomic position and environmental conditions by subjective cognitive decline among LGBTQIA+ adults aged 50 years or older in California.

|  |  | Subjective cognitive decline |  |  |  |
|---|---|---|---|---|---|
|  | Overall | No | Yes |  |  |
|  | n = 3869 | n = 3301 | n = 568 |  |  |
|  | n (%) | n (%) | n (%) | *P* | Missing |
| Education levels |  |  |  | 0.002 | 0.3% |
| High school diploma/GED or less | 102 (2.6) | 79 (2.4) | 23 (4.0) |  |  |
| Some college | 932 (24.1) | 768 (23.3) | 164 (28.9) |  |  |
| 4-year degree | 1201 (31.0) | 1027 (31.1) | 174 (30.6) |  |  |
| Graduate degree | 1624 (42.0) | 1418 (43.0) | 206 (36.3) |  |  |
| Unemployment | 385 (10.0) | 285 (8.6) | 100 (17.6) | <0.001 | 0.1% |
| Household income |  |  |  | <0.001 | 2.2% |
| \$0-\$30,000 | 655 (16.9) | 492 (14.9) | 163 (28.7) |  |  |
| \$30,001-\$60,000 | 707 (18.3) | 581 (17.6) | 126 (22.2) |  |  |
| \$60,001-\$90,000 | 602 (15.6) | 519 (15.7) | 83 (14.6) |  |  |
| \$90,001-\$120,000 | 572 (14.8) | 517 (15.7) | 55 (9.7) |  |  |
| ≥\$120,001 | 1246 (32.2) | 1120 (33.9) | 126 (22.2) |  |  |
| Household assets |  |  |  | <0.001 | 12.0% |
| \$0-\$9,999 | 502 (13.0) | 382 (11.6) | 120 (21.1) |  |  |
| \$10,000-\$100,000 | 606 (15.7) | 484 (14.7) | 122 (21.5) |  |  |
| \$100,001-\$500,000 | 700 (18.1) | 607 (18.4) | 93 (16.4) |  |  |
| ≥\$500,001 | 1597 (41.3) | 1427 (43.2) | 170 (29.9) |  |  |
| Financial insecurity | 1001 (25.9) | 765 (23.2) | 236 (41.5) | <0.001 | 0.5% |
| Food insecurity | 1352 (34.9) | 1052 (31.9) | 300 (52.8) | <0.001 | 0.1% |
| Unstably housed | 500 (12.9) | 375 (11.4) | 125 (22.0) | <0.001 | 0.4% |
| Relies on public transportation | 1314 (34.0) | 1092 (33.1) | 222 (39.1) | 0.02 | 1.6% |
| LGBTQIA+ community safety |  |  |  | <0.001 | 9.4% |
| Not safe | 803 (20.8) | 627 (19.0) | 176 (31.0) |  |  |
| Safe for SM only | 649 (16.8) | 551 (16.7) | 98 (17.3) |  |  |
| Safe for GM only | 69 (1.8) | 55 (1.7) | 14 (2.5) |  |  |
| Safe for both groups | 1986 (51.3) | 1754 (53.1) | 232 (40.8) |  |  |

GED, general educational development; GM, gender minority; LGBTQIA+, lesbian, bisexual, gay, transgender, queer, intersex, asexual and other sexual or gender minority; SM, sexual minority

Overall, 14.7% of participants reported experiencing SCD. Compared to participants who did not report SCD, those reporting SCD were more likely to be aged 70 years or older (38.7% vs. 33.2%, p=0.04), gender diverse assigned male at birth (5.6% vs. 2.6%, p=0.004), bisexual or pansexual individuals (11.8% vs. 8.7%, p=0.02), to have prior military service (12% vs. 9.1%, p=0.04), and to be currently living with HIV (23.2% vs. 15.1%, p<0.001).

Pairwise correlations of each socioeconomic and contextual factor of interest are shown in **Figure 1**. Education levels showed positive correlations with household income (r=0.32, p<0.001) and assets (r=0.29, p<0.001), while negatively correlating with financial insecurity (r=-0.23, p<0.001) and food insecurity (r=-0.26, p<0.001). Household income and assets were moderately positively correlated (r=0.54, p<0.001) and both negatively correlated with financial insecurity, food insecurity, and housing instability (all p<0.001). Financial and food insecurity were strongly correlated (r=0.65, p<0.001) and both positively associated with housing instability (r=0.35 and r=0.36, respectively, both p<0.001). Lack of LGBTQIA+ community safety was positively correlated with financial insecurity (r=0.19, p<0.001), food insecurity (r=0.23, p<0.001), and housing instability (r=0.15, p<0.001). Reliance on public transportation showed weak but significant positive correlations with various insecurity measures.

**Figure 1.**
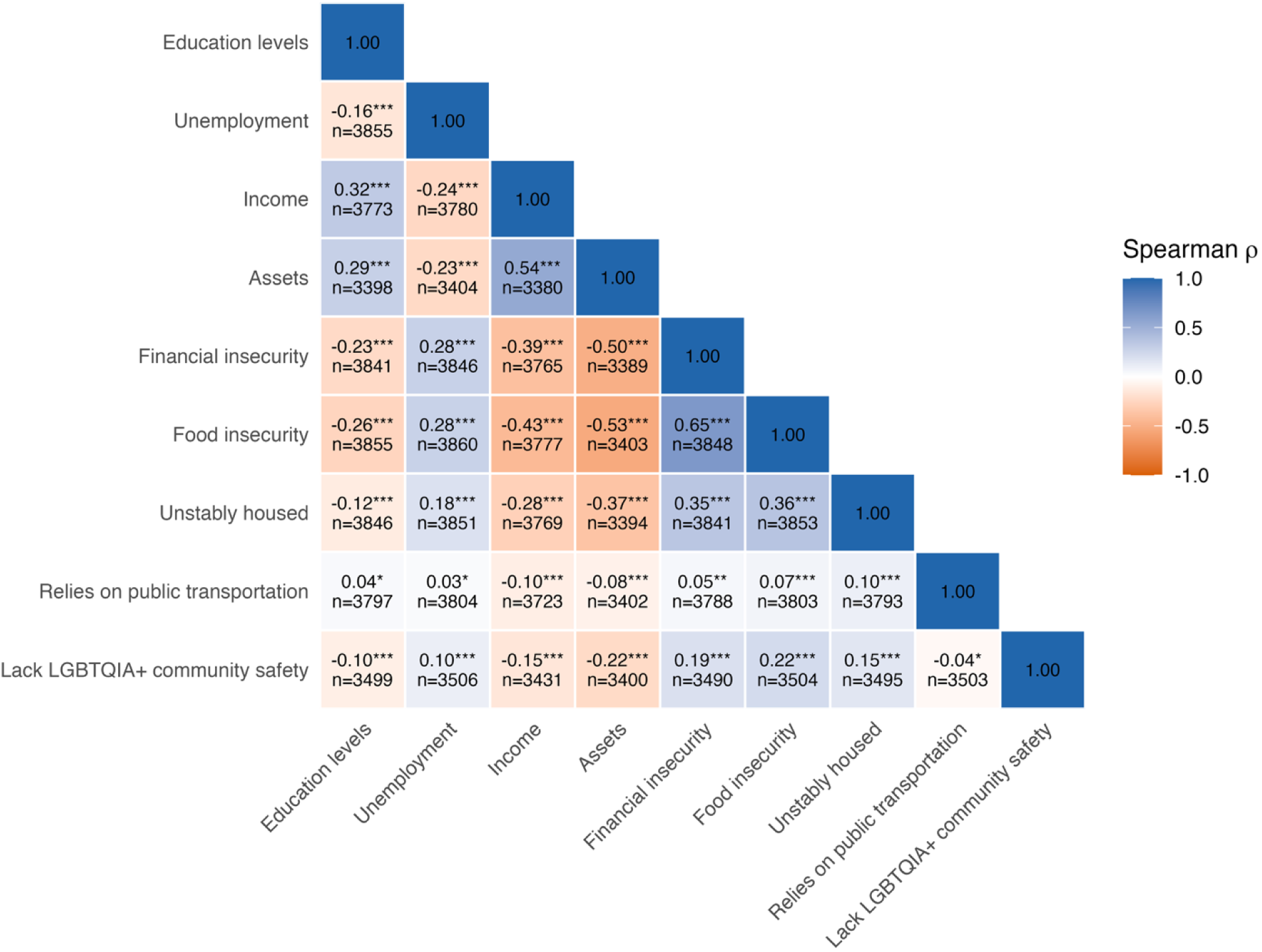
Spearman correlation and sample size between socioeconomic position and environmental conditions among LGBTQIA+ adults aged 50 years or older in California. *** *P* < 0.001; ** *P* < 0.01; * *P* < 0.05.

### Regression modeling

In unadjusted and adjusted models, nearly all socioeconomic and socioenvironmental conditions were significantly associated with SCD (**Figure 2** and **Supplemental Table 1**). In models that regressed SCD on education, household income, household assets, being in a lower position for any of these socioeconomic factors was associated with higher odds of SCD, with progressively stronger associations at lower socioeconomic positions. For example, in adjusted models, respondents with the lowest household income and assets had 2.03 (95% CI 1.25, 3.31) times and 2.55 (95% CI 1.96, 3.32) times higher odds of SCD compared to the highest income and asset groups. At moderate household income ($30,000-$60,000) and assets ($10,000-$100,000), respondents had 1.76 (95% CI 1.34, 2.31) times and 2.02 (95% CI 1.56, 2.62) times higher odds of SCD compared to the highest income and asset groups.

**Figure 2.**
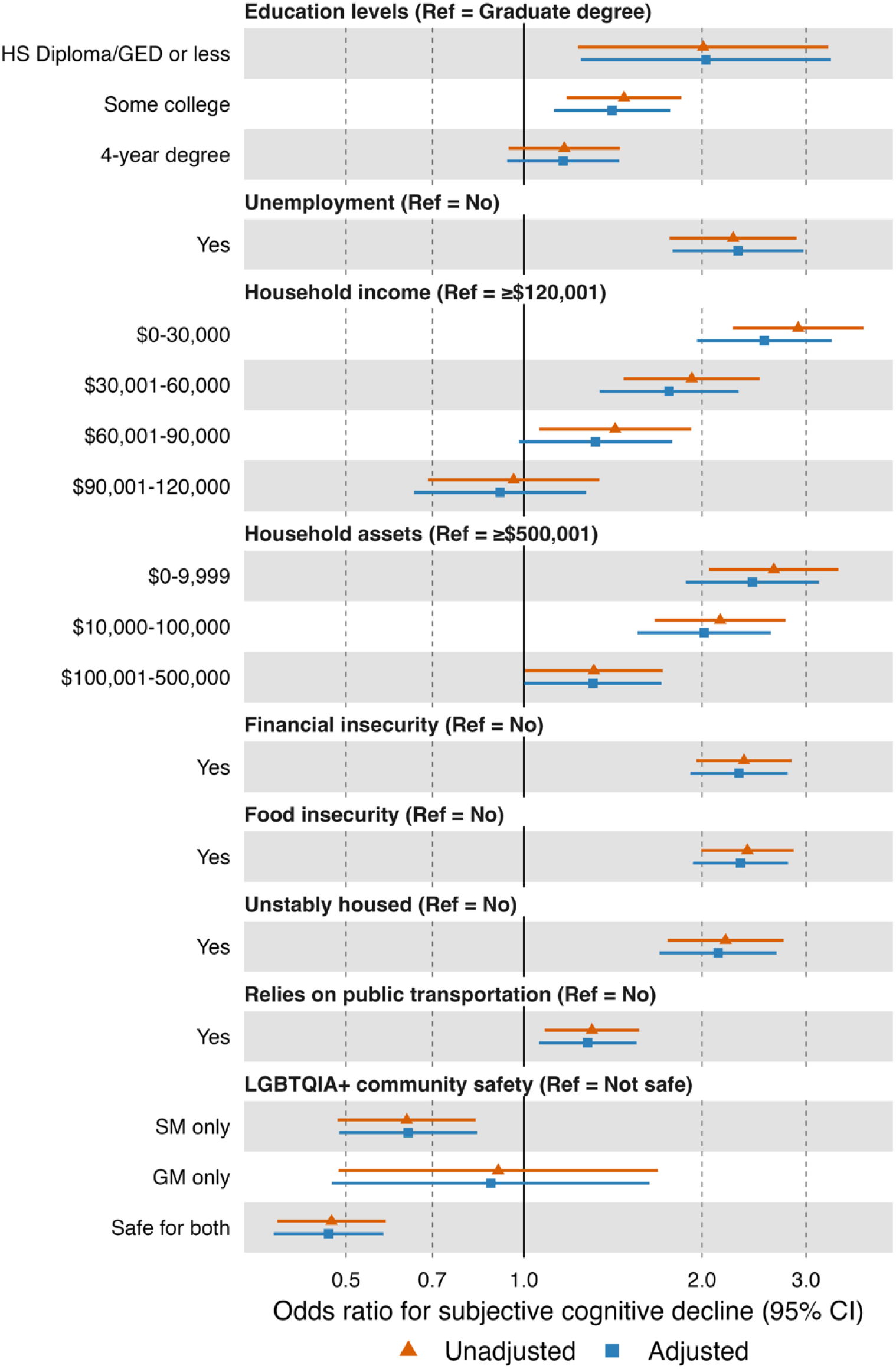
Associations of indicators of socioeconomic position and environmental conditions with subjective cognitive decline among LGBTQIA+ adults aged 50 years or older in California. Odds ratios from logistic models are shown. Adjusted models accounted for age groups, gender groups, ethnoracial groups, US birth, urban residence, military service, hearing or vision loss, history of trauma, social support, caregiving, and living with HIV. Coefficients are shown in Supplemental Table 1. CI, confidence interval; HS, high school, GED, general educational development; GM, gender minority, SM, sexual minority.

Similarly, unemployment, financial insecurity, food insecurity, unstable housing, reliance on public transportation were all associated with higher odds of SCD, with adjusted ORs ranging from 1.28 to 2.30. Regarding LGBTQIA+ community safety, those who reported their community as safe for both sexual and gender minorities had lower odds of SCD (adjusted OR 0.47; 95% CI 0.38, 0.58) compared to those who felt their community was not safe. In sensitivity analysis that additionally adjusting for mental health symptoms and substance use, we observed attenuation of the estimated associations (**Supplemental Table 1**). Several associations remained statistically significant (*e.g.*, education, household income, financial and food insecurity, and perceived LGBTQIA+ community safety), whereas others (*e.g.*, unstable housing and reliance on public transportation) were reduced in magnitude and their 95% CIs included the null.

### Dominance analysis

Estimates of the multivariable logistic model that included all socioeconomic and socioenvironmental conditions while controlling for potential confounders, as outlined in our main analysis, are shown in **Supplemental Table 2**; results from complete cases and multiply imputed data indicated minimal differences. The dominance analysis suggested that among midlife and older LGBTQIA+ respondents, household assets were the most important factor influencing SCD, explaining 4.4% of the variance (95% CI 3.5%, 5.4%; **Figure 3**). This was followed closely by household income, which explained 2.9% of the variance (95% CI 2.2%, 3.8%), and perceived community safety, which explained 2.2% of the variance (95% CI 1.4%, 3.1%). Unemployment, financial insecurity, housing instability, food insecurity, education levels, and reliance on public transit each explained <1% of the variance in SCD among respondents. Sensitivity analyses that additionally accounted for mental health symptoms and substance use in dominance analysis showed attenuation in the variance explained in SCD by each factor (**Supplemental Figure 2**). Reduction in the variance explained in SCD for assets, household income, and perceived community safety were all by at least 1 percentage point.

**Figure 3.**
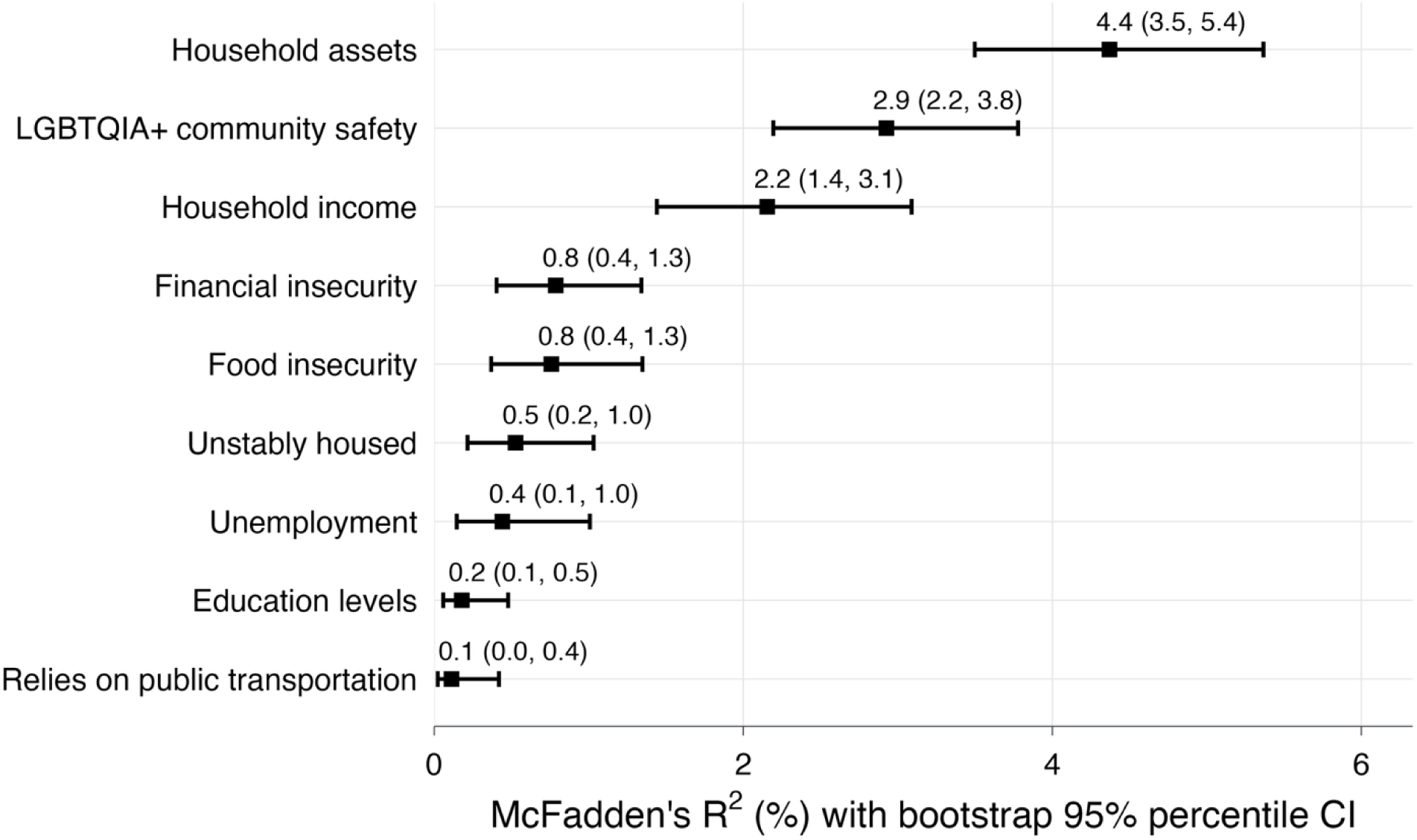
General dominance estimates (using McFadden’s R^2^) and 95% percentile confidence intervals over 1000 bootstrap resamples for socioeconomic and environment factors in relation to subjective cognitive decline among LGBTQIA+ adults aged 50 years or older in California.z

## DISCUSSION

Consistent with prior studies among midlife and older LGBTQIA+ adults from population-based samples (Cicero et al., 2023; Flatt et al., 2021; Fredriksen-Goldsen et al., 2022; Hsieh et al., 2021), nearly 15% of midlife and older LGBTQIA+ respondents in this California sample reported SCD with higher percentages reported in certain subgroups such as bisexual and pansexual people as well as gender diverse people assigned male at birth. Midlife and older LGBTQIA+ respondents with SCD were more likely to experience social and economic disadvantage compared to those without SCD, even after accounting for age, gender, ethnoracial groups, US birth, urban residence, prior military service, and HIV status. The most important socioeconomic and socioenvironmental contributors to SCD were household assets (explaining 4.4% of the variance), followed by household income (2.9% of the variance) and community safety for LGBTQIA+ people (2.2% of the variance). Other factors, such as unemployment, financial insecurity, housing instability, food insecurity, and education level, were found to be less important, with each factor explaining less than 1% of the variance in SCD. Because our sample was predominantly cisgender and gay or lesbian, and include relatively more socially and economically advantaged respondents, these aggregate estimates may primarily reflect the experiences of these groups and mask subgroup differences in association of economic and social conditions with SCD for transgender and diverse people, bisexual people, and racially minoritized LGBTQIA+ people. Thus, the observed associations may underestimate the potential magnitude of social and economic disparities in SCD among midlife and older LGBTQIA+ adults.

Our result is consistent with the broader literature on social factors and SCD among the general population (Singh-Manoux et al., 2005; Lövdén et al., 2020; Luo et al., 2025) and on research focusing on the economic marginalization LGBTQIA+ individuals experience and the proposed biological pathways between minority stress and physical health (Correro II & Nielson, 2020; Flentje et al., 2020, 2022; Hendricks & Testa, 2012; Kinitz et al., 2025; Meyer, 2003). Chronic minority stress may be related to epigenetic changes and alterations in the hypothalamic-pituitary-adrenal axis functioning associated with accelerated cognitive decline in LGBTQIA+ individuals (Correro II & Nielson, 2020; Flentje et al., 2020; Hatzenbuehler & McLaughlin, 2014). Complementary work has documented a higher prevalence of SCD among LGBTQIA+ people, identified social isolation and depressive symptoms as proximal mediators of cognitive decline in this population (Correro II & Nielson, 2020; Fredriksen-Goldsen et al., 2018; Hsieh et al., 2021; Kim et al., 2025), and highlighted poorer economic conditions as a potentially important socially patterned exposure that may result from minority stress (Kinitz et al., 2025; Kinitz, Tran, Shahidi, et al., 2026; Kinitz, Tran, Soltani, et al., 2026; Nguyen et al., 2024). Notably, in our study, associations between lower socioeconomic position and SCD remained robust after adjusting for mental health symptoms and substance use, though we observed attenuation of the explained SCD variance after accounting for mental health and substance use in the dominance analysis. This suggests the potential importance of these factors for SCD in LGBTQIA+ individuals and highlight the need for future longitudinal research to better understand the mechanism between economic marginalization, mental health and substance use, and later-life cognitive decline.

An important contribution of our study is the identification of community safety as a socioenvironmental correlate of cognitive health in LGBTQIA+ populations. Respondents who perceived their community as safe for sexual and/or gender minority individuals had 53% lower odds of reporting SCD. Although research directly examining the association of community safety and SCD among LGBTQIA+ is limited, this result aligns with evidence examining the association of perceived community safety and physical health (Flentje et al., 2022). Relatedly, area-based characteristics that capture aspects of community safety, such as place-based disorder (*e.g.*, signs of neglect, violence, or lack of perceived safety), have been associated with higher stress perceptions among LGBTQIA+ adults and greater depressive symptoms among transgender and gender diverse adults (Tran et al., 2026). These findings underscore the broader influence of the social environment on cognitive health in LGBTQIA+ populations and highlight the need for further research into the mechanisms through which community safety may shape SCD.

Results from dominance analysis further suggest that both financial security across the life course and the broader social environments may be the most relevant socioeconomic and socioenvironmental condition to SCD. Household assets, which represent accumulated wealth over the life course (Krieger et al., 1997), were shown to explain the most variance in SCD. One’s assets (or lack thereof) may reflect the long-term consequences of earlier financial strains that leave LGBTQIA+ people unable to build their savings or possess the necessary financial resources to support cognitive aging and care in later life. This is consistent with work suggesting that LGBTQIA+ individuals experience heightened employment precarity and economic marginalization related to minority stress (Kinitz et al., 2025; Nguyen et al., 2024), which may accumulate over time to shape cognitive health. Because assets reflect long-term financial trajectories, they may be difficult to modify later in life, particularly for economically marginalized groups. This highlights the potential value of future research focusing on interventions that promote financial security earlier in the life course, such as stable income opportunities and retirement and financial planning programs tailored to the needs of LGBTQIA+ communities. By contrast, household income and perceived community safety highlight the potential relevance of policies and programs that address the current social and economic circumstances for midlife and older LGBTQIA+ people, especially those with SCD. Such efforts may include addressing economic marginalization that LGBTQIA+ people face, such as unemployment, poorer job quality, and gender-based discrimination (Fredriksen Goldsen et al., 2022; Kinitz et al., 2025; Nguyen et al., 2024), which may contribute to fewer financial resources that would allow LGBTQIA+ people to live in communities with more supportive resources.

### Limitations

This study has several limitations. First, the cross-sectional design precludes causal interpretation, as cognitive impairment in older age may itself contribute to greater financial insecurity and poorer living conditions. Second, the outcome was self-reported. Prior work has noted that SCD is an imperfect indicator of objective cognitive impairment (Romanelli et al., 2024), and future studies would benefit from incorporating cognitive testing to better understand how socioeconomic and socioenvironmental conditions relate to cognitive decline. Relatedly, the cross-sectional design does not allow us to examine changes in cognition over time, which may be more biologically relevant for understanding the influence of social and economic factors.

Third, selection bias is possible if LGBTQIA+ individuals experiencing greater economic marginalization and SCD were less likely to participate. Fourth, our analyses assume no model misspecification and that included covariates were sufficient to control for confounding, though unmeasured confounding may be present. Fifth, while minimal differences were observed between the multiple imputation and complete-case analysis, these methods may still be biased under missing-not-at-random or misspecification of the imputation model. Sixth, participants included in this study were derived from a convenience, with an overrepresentation of White, cisgender, and gay/lesbian respondents and may not be representative of LGBTQIA+ individuals in California. Furthermore, while the survey was offered in four languages, nearly all participants completed it in English. These factors limit the generalizability of our findings.

Finally, our analyses considered LGBTQIA+ adults as an aggregate population and were not designed to examine heterogeneity in these associations across sexual orientation, gender identity, or intersecting social identities. Prior research suggests that experiences of SCD may differ across sexual and gender minority subgroups and at the intersections of gender and ethnoracial identity (Cicero et al., 2023; Flatt et al., 2021). Future studies with larger samples of transgender, gender-diverse, and other underrepresented LGBTQIA+ populations should examine whether socioeconomic and socioenvironmental correlates of SCD differ across these groups.

## CONCLUSION

Notwithstanding these limitations, the current analysis contributes to the literature on aging and LGBTQIA+ health by offering new evidence on the relative importance of social and economic conditions that may support cognitive aging in midlife and older LGBTQIA+ adults. Using regression and dominance analysis, our study identified both economic security and community safety as factors that can both inform future research and guide focused interventions. Future research examining interventions to improve financial literacy and retirement planning may help support LGBTQIA+ people as they age. Other work might consider the potential role of social safety net programs, such as Social Security and Supplemental Security Income, in mitigating the economic marginalization that LGBTQIA+ people experience across the life course.

## Data Availability

All reproducible code for the analysis can be found at: https://github.com/ntran756/cda_older_adults.git. Requests for data access should be made to the California Department of Aging.

## Acknowledgements

We would like to acknowledge the efforts of the LGBTQIA+ older adults who participated in this study, the community organizations across California that reached participants, and the study collaborators: University of California, Berkeley, Citris and the Banatao Institute and School of Social Welfare, Center for Advanced Aging Services and Openhouse.

## Disclaimer

N/A

## Code and Data Availability Statement

All reproducible code for the analysis can be found at: https://doi.org/10.5281/zenodo.24120124. Requests for data access should be made to the California Department of Aging.

