## Supplemental for "Socioeconomic position, socioenvironmental conditions, and subjective cognitive decline among midlife and older LGBTQIA+ adults in California"

### Supplemental Materials

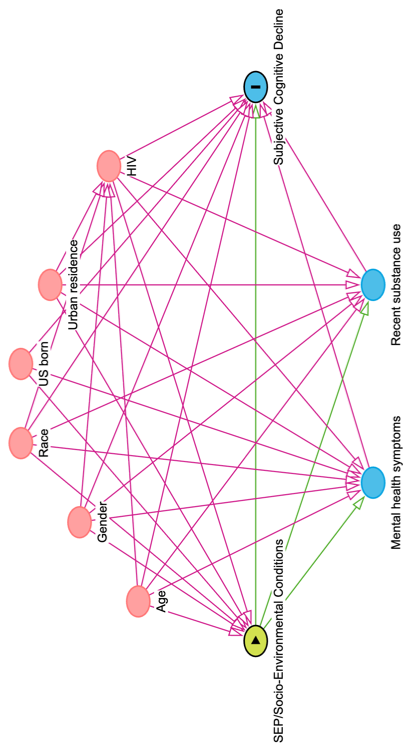

**Supplemental Figure 1.** Directed acyclic graph showing the hypothesized relationships of the exposure-outcome relationship for socioeconomic positions (SEP), environmental conditions, and subjective cognitive decline.

Red circles indicate potential confounders or variables that are associated with the exposures and outcome and were assumed to have occurred prior to the exposure. Red arrows indicate biasing pathways. Green arrows indicate causal pathways. Mental health symptoms and recent substance use are assumed to be mediators and lie along the causal pathways between the exposure and outcome.

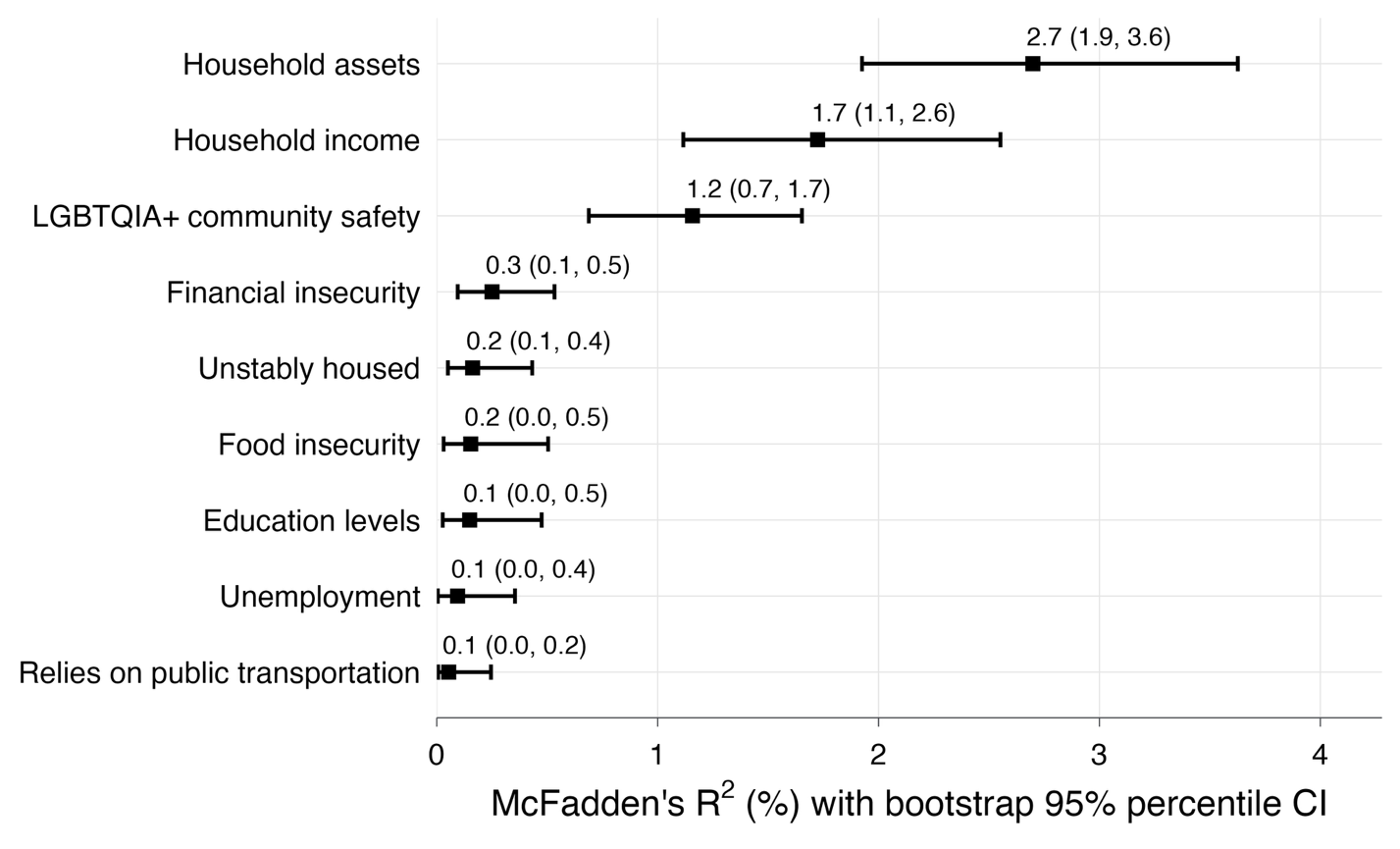

**Supplemental Figure 2**. General dominance estimates (using McFadden’s R^2^) and 95% percentile confidence intervals over 1000 bootstrap resamples for socioeconomic and environment factors in relation to subjective cognitive decline that additionally adjusted for mental health symptoms (loneliness, depressive symptoms, posttraumatic stress disorder symptoms) and substance use (current cigarette smoker and alcohol use) among LGBTQIA+ adults aged 50 years or older in California.

**Supplemental Table 1.** Odds ratios and 95% confidence intervals for associations of socioeconomic and environmental factors with subjective cognitive decline among LGBTQIA+ adults aged 50 years or older in California.

|  | | Main Analysis | | Sensitivity Analysis |
| --- | --- | --- | --- | --- |
| Characteristics | | Unadjusted Odds Ratio (95% Confidence Interval) | Adjusted Odds Ratio (95% Confidence Interval)^a^ | Adjusted Odds Ratio (95% Confidence Interval)^b^ |
| Education levels | |  |  |  |
|  | High school diploma/GED or less | 2.01 (1.24, 3.27) | 2.03 (1.25, 3.31) | 1.67 (1.05, 2.65) |
|  | Some college | 1.47 (1.18, 1.84) | 1.41 (1.12, 1.77) | 1.12 (0.90, 1.40) |
|  | 4-year degree | 1.17 (0.94, 1.45) | 1.16 (0.94, 1.45) | 1.06 (0.86, 1.30) |
| Unemployment | | 2.26 (1.76, 2.89) | 2.30 (1.78, 2.97) | 1.31 (1.01, 1.70) |
| Household income | |  |  |  |
|  | $0-30,000 | 2.90 (2.25, 3.74) | 2.55 (1.96, 3.32) | 1.48 (1.13, 1.94) |
|  | $30,001-60,000 | 1.93 (1.47, 2.51) | 1.76 (1.34, 2.31) | 1.22 (0.94, 1.60) |
|  | $60,001-90,000 | 1.42 (1.06, 1.91) | 1.32 (0.98, 1.78) | 1.04 (0.78, 1.39) |
|  | $90,001-120,000 | 0.95 (0.68, 1.32) | 0.91 (0.65, 1.27) | 0.82 (0.59, 1.12) |
| Household assets | |  |  |  |
|  | $0-9,999 | 2.67 (2.07, 3.44) | 2.43 (1.88, 3.16) | 1.38 (1.06, 1.81) |
|  | $10,000-100,000 | 2.13 (1.66, 2.73) | 2.02 (1.56, 2.62) | 1.36 (1.05, 1.76) |
|  | $100,001-500,000 | 1.33 (1.01, 1.74) | 1.31 (1.00, 1.71) | 1.03 (0.80, 1.34) |
| Financial insecurity | | 2.36 (1.96, 2.84) | 2.31 (1.91, 2.79) | 1.43 (1.18, 1.74) |
| Food insecurity | | 2.39 (1.99, 2.86) | 2.32 (1.93, 2.80) | 1.44 (1.18, 1.74) |
| Unstably housed | | 2.19 (1.75, 2.75) | 2.13 (1.70, 2.68) | 1.21 (0.96, 1.53) |
| Relies on public transportation | | 1.30 (1.08, 1.57) | 1.28 (1.06, 1.55) | 1.15 (0.96, 1.39) |
| LGBTQIA+ community safety | |  |  |  |
|  | Safe for SM only | 0.63 (0.48, 0.83) | 0.64 (0.49, 0.83) | 0.90 (0.69, 1.17) |
|  | Safe for GM only | 0.90 (0.48, 1.69) | 0.88 (0.47, 1.63) | 0.92 (0.51, 1.66) |
|  | Safe for both groups | 0.47 (0.38, 0.59) | 0.47 (0.38, 0.58) | 0.73 (0.59, 0.91) |

AUDIT-C, Alcohol Use Disorders Identification Test-Consumption; CI, confidence interval; GM, gender minority OR, odds ratio; PCL-6, 6-item Posttraumatic Stress Disorder Checklist; and PHQ-2, 2-item Patient Health Questionnaire; SM, sexual minority; UCLA, University of California Los Angeles.

^a^ Model adjusted for age groups, gender groups, ethnoracial groups, being US born, urban residence, military service, and living with HIV.

^b^ Model from sensitivity analysis further adjusted for current cigarette smoking, AUDIT-C scores for alcohol use disorder, UCLA 3-item score for loneliness, PHQ-2 score for depressive symptoms, and PCL-6 scores for posttraumatic stress disorder symptoms in addition to the abovementioned covariates.

**Supplemental Table 2.** Full model results of multivariable logistic regression from complete case analysis and multiple imputation including all socioeconomic and environmental factors while controlling for potential confounders among LGBTQIA+ adults aged 50 years or older in California.

|  |  | Complete Case | Multiple Imputation |
| --- | --- | --- | --- |
|  | | Odds Ratio  (95% Confidence Interval) | Odds Ratio  (95% Confidence Interval) |
| Education levels | |  |  |
|  | HS diploma/GED or less | 1.18 (0.63, 2.12) | 1.24 (0.73, 2.10) |
|  | Some college | 0.99 (0.75, 1.30) | 0.97 (0.75, 1.24) |
|  | 4-year degree | 1.01 (0.79, 1.30) | 1.01 (0.80, 1.27) |
| Unemployment | | 1.45 (1.05, 1.99) | 1.47 (1.10, 1.96) |
| Household income | |  |  |
|  | $0-30,000 | 1.17 (0.81, 1.70) | 1.29 (0.91, 1.81) |
|  | $30,0001-60,000 | 1.13 (0.81, 1.58) | 1.15 (0.84, 1.57) |
|  | $60,001-90,000 | 0.95 (0.67, 1.34) | 1.03 (0.75, 1.41) |
|  | $90,001-120,000 | 0.71 (0.48, 1.02) | 0.79 (0.56, 1.12) |
| Household income | |  |  |
|  | $0-9,999 | 1.08 (0.74, 1.57) | 1.05 (0.73, 1.51) |
|  | $10,000-100,000 | 1.17 (0.84, 1.62) | 1.16 (0.84, 1.60) |
|  | $100,001-500,000 | 1.01 (0.74, 1.36) | 1.02 (0.76, 1.36) |
| Financial insecurity | | 1.21 (0.90, 1.63) | 1.34 (1.02, 1.75) |
| Food insecurity | | 1.38 (1.02, 1.85) | 1.36 (1.03, 1.78) |
| Unstably housed | | 1.41 (1.06, 1.87) | 1.26 (0.97, 1.64) |
| Relies on public transportation | | 1.16 (0.93, 1.44) | 1.20 (0.98, 1.47) |
| LGBTQIA+ community safety | |  |  |
|  | Safe for SM only | 0.78 (0.57, 1.05) | 0.77 (0.58, 1.02) |
|  | Safe for GM only | 1.05 (0.53, 1.96) | 0.89 (0.47, 1.70) |
|  | Safe for both | 0.62 (0.48, 0.79) | 0.59 (0.47, 0.75) |
| Age groups | |  |  |
|  | 60-69 | 1.04 (0.79, 1.37) | 1.11 (0.87, 1.43) |
|  | ≥70 | 1.52 (1.13, 2.05) | 1.69 (1.29, 2.23) |
| Minoritized ethnoracial group | | 0.84 (0.64, 1.08) | 0.90 (0.71, 1.14) |
| Gender groups | |  |  |
|  | Cisgender woman | 1.53 (1.19, 1.98) | 1.45 (1.14, 1.83) |
|  | Gender diverse AFAB | 1.31 (0.80, 2.10) | 1.18 (0.76, 1.84) |
|  | Gender diverse AMAB | 1.97 (1.17, 3.23) | 1.97 (1.26, 3.09) |
|  | Transgender man | 1.17 (0.34, 3.12) | 1.80 (0.80, 4.05) |
|  | Transgender woman | 1.79 (0.88, 3.40) | 1.65 (0.87, 3.12) |
| US born | | 1.04 (0.69, 1.62) | 0.98 (0.67, 1.44) |
| Urban residence | | 1.03 (0.79, 1.35) | 1.12 (0.87, 1.43) |
| Military service | | 1.47 (1.06, 2.01) | 1.32 (0.98, 1.77) |
| Living with HIV | | 1.91 (1.45, 2.50) | 1.73 (1.34, 2.24) |
| No. observations | | 3205 | 3869 |

GM, gender minority; SM, sexual minority.
